# Do Large Language Models Read or Remember? Analyzing LLM Performance in Biomedical Text Mining With Progressive Content Removal and Counterfactual Results

**DOI:** 10.64898/2026.02.02.26345347

**Authors:** Paul Windisch, Carole Koechli, Fabio Dennstädt, Daniel M. Aebersold, Daniel R. Zwahlen, Robert Förster, Christina Schröder

**Affiliations:** Department of Radiation Oncology, Cantonal Hospital Winterthur, Winterthur, Switzerland; Department of Radiation Oncology, Inselspital, Bern University Hospital, University of Bern, Bern, Switzerland

**Keywords:** Natural language processing, Large language models, Memory, Reasoning models

## Abstract

**Purpose:** Large language models (LLMs) can classify biomedical documents accurately, but strong performance does not prove they are using the supplied text rather than identifier-triggered parametric knowledge. We tested whether oncology trial-success classification reflects “reading” of abstract evidence or “remembering” of known trials.

**Methods:** We used a corpus of 250 two-arm oncology randomized controlled trials from seven major journals (2005 - 2023) and asked the flagship models of three commercial vendors (OpenAI, Google, and Anthropic) to output a single label indicating whether the primary endpoint was met. For each trial we created five deterministic inputs: title+abstract (baseline), title-only, DOI-only, counterfactual title+abstract with the primary endpoint outcome minimally flipped, and the same counterfactual title+abstract paired with the original DOI to induce an identifier-text conflict.

**Results:** With full title+abstract, models achieved near-ceiling performance (accuracy and F1 Score 0.96 - 0.97) and high format adherence (97.2 - 100%). Performance degraded stepwise with content removal (title-only accuracy and F1 Score 0.79 - 0.88, DOI-only 0.63 - 0.67), consistent with above-chance identifier-driven signal. Under counterfactual results, models followed the edited evidence (accuracy and F1 Score 0.96 - 0.99 against inverted labels). Adding the real DOI minimally affected GPT (accuracy and F1 Score ≈ 0.99) but modestly reduced Gemini (accuracy and F1 Score ≈ 0.97) and Claude (accuracy and F1 Score ≈ 0.95), mainly via lower sensitivity.

**Conclusion:** LLMs robustly track explicit endpoint statements in abstracts, yet identifiers can support above-chance predictions and occasionally compete with textual evidence. Progressive ablations plus counterfactual conflicts provide a practical, reproducible audit for grounding in biomedical LLM evaluations.

## Introduction

Large language models (LLMs) are increasingly used for biomedical text classification and information extraction, including clinical trial matching, automated abstraction from electronic health records, and literature screening in evidence synthesis workflows.^1–3^

However, LLM success on biomedical text tasks is not, by itself, evidence that the model is reading and reasoning over the provided input. A defining feature of modern foundation models is that they store substantial “parametric knowledge” acquired during pretraining, and in some settings they can behave like implicit knowledge bases that retrieve facts from model weights rather than from the prompt context.^4^ In medicine specifically, large models have been shown to encode clinically relevant knowledge and to perform strongly on medical question-answering benchmarks, reinforcing that model parameters can serve as a powerful source of prior information independent of any supplied document.^5^ For biomedical publishing and evidence synthesis, this distinction is consequential: users may expect the system to follow the text in front of it, while the model may instead answer using remembered associations with the underlying paper, trial name, or venue.

Concerns about “remembering” are amplified by two related phenomena: memorization and data contamination. Previous work has demonstrated that large language models can leak or reproduce training data under targeted querying, indicating that verbatim or near-verbatim sequences can be memorized and later elicited.^6^ More recent work has developed formal measures for quantifying extractability and memorization risk, highlighting that the phenomenon can be measured and varies with model and inference choices.^7^ Separately, systematic analyses have documented that benchmark contamination is widespread in the LLM era and can occur at non-trivial rates, creating the possibility that models appear to “solve” tasks by recognizing previously seen evaluation items rather than generalizing.^8^ In biomedical domains, where many inputs (titles, abstracts, DOIs) are publicly available and plausibly included in pretraining corpora, a model might correctly predict a randomized controlled trial’s (RCT) outcome without genuinely using the evidence contained in the abstract.

A growing methodological literature has therefore begun to explicitly separate contextual evidence use from prior (parametric) knowledge. Work on knowledge conflicts shows that language models do not integrate context and prior knowledge uniformly, and may privilege prior knowledge when the model is “familiar” with the entity or topic referenced.^9^ Methods such as contrastive decoding have been proposed to reduce overreliance on encoded priors and improve the use of contextual information when the prompt provides relevant evidence.^10^ Relatedly, counterfactual paradigms have been used to disentangle parametric from contextual knowledge by constructing prompts in which the provided context is intentionally altered, enabling direct tests of whether a model follows the prompt or defaults to internal memory.^11^ In the biomedical setting, dedicated benchmarks have further emphasized that knowledge conflicts and unfaithful grounding are practical risks when users provide incomplete, contradictory, or incorrect context.^12^ Despite these advances, biomedical evaluations that use real documents rarely operationalize a direct, auditably reproducible “read-versus-remember” test that isolates the informational contribution of the input text from that of identifiers.

This paper focuses on that methodological gap using the concrete task of classifying oncology RCTs as positive versus negative with respect to primary endpoint attainment. To distinguish input-grounded “skill” from identifier-driven “memory/recognition,” we leverage an evaluation framework based on progressive content removal and counterfactual results. The key intuition is diagnostic: If performance remains meaningfully above chance when only identifiers are provided, this indicates that the model is able to remember the contents of the trial and abstract. If predictions then fail to flip under counterfactual results text, then the model’s apparent “understanding” is more consistent with recognition or memory dominance than with faithful reading of the supplied evidence.

We evaluate three widely used commercial model families under default settings to reflect typical end-user deployment conditions. By combining progressive ablations with counterfactual conflict tests, this study aims to provide an empirically grounded answer to a practical question that is often implicit in biomedical LLM benchmarking: when an LLM appears to “read” a clinical trial abstract correctly, is it truly reading - or is it remembering?

## Methods

This study evaluates whether LLM performance on biomedical trial-success classification is primarily driven by the information contained in the provided input text (“reading”) or by recognition and parametric knowledge associated with identifiers (“remembering”). We used an existing, previously published dataset from the same author group consisting of 250 RCTs drawn from seven major medical journals (British Medical Journal, JAMA, JAMA Oncology, Journal of Clinical Oncology, Lancet, Lancet Oncology, and New England Journal of Medicine) published between 2005 and 2023.^13^ In the original publication, eligible trials were restricted to designs with exactly two arms and a single primary endpoint, and abstracts were retrieved via PubMed and parsed from text to create the study corpus. For the present analysis, we reused the released dataset and corresponding ground-truth labels.

Ground-truth labels were adopted from the original dual-annotation procedure performed by two authors (P.W., C.K.), in which trials were classified as positive if the primary endpoint was met and negative otherwise. The annotation workflow included an initial calibration phase, followed by independent labeling and consensus resolution of discrepancies. Full texts or protocols were consulted only when the abstract did not clearly report the primary endpoint and its results. We did not modify labels or repeat manual annotation for the present analysis.

To distinguish evidence use from identifier-driven recognition, we generated five input variants for each RCT. The baseline condition contained the trial title and abstract (title + abstract). A title-only condition contained only the trial title. A DOI-only condition contained only the DOI string. A counterfactual “fake-results” condition contained the potentially edited title and an edited abstract in which the reported primary endpoint outcome was flipped (positive to negative or negative to positive) while keeping all other content maximally unchanged. Counterfactual abstracts were created by identifying the sentence(s) reporting primary endpoint attainment (typically in the results and/or conclusion) and minimally editing the outcome language (e.g., reversing whether the primary endpoint was met, or reversing the direction of statistical significance statements tied to the primary endpoint). Background, design, population, interventions, and secondary outcome text were left unchanged unless modification was required to maintain internal coherence. The original and counterfactual texts are provided in the repository. Lastly, we sent the counterfactual title and abstract together with the original DOI to try to “bait” the model into relying on its parametric knowledge and ignoring the actual text in front of it.

The task for the LLMs was to determine, from the provided information, whether the trial met its primary endpoint. To align with the previously published workflow and minimize ambiguity, we used an explicit instruction format requiring the model to output exactly one token-level label: POSITIVE or NEGATIVE in all caps.

The following system prompt was used: “You will be provided with information about a randomized controlled oncology trial. Your task will be to classify if the trial was positive, i.e. if it met its primary endpoint, or negative, i.e. if it did not meet its primary endpoint. Your response should be either the word POSITIVE (in all caps) or NEGATIVE (in all caps). Do not output anything else” The user prompt consisted of the respective input variant (title + abstract, title-only, DOI-only, counterfactual title + abstract, DOI and counterfactual title + abstract).

Three commercial LLM families were evaluated via their vendor APIs in a local pipeline (Claude, Gemini, and GPT). Models were queried under default vendor settings to reflect typical end-user deployment conditions. No additional decoding parameters were set and no seed was specified. We used the model snapshots *gpt-5.2-2025-12-11, gemini-3-flash-preview*, and *claude-opus-4-5-20251101*.

Responses were considered valid only if the returned text matched exactly POSITIVE or NEGATIVE after trimming whitespace. Any other output was recorded as invalid for downstream summaries.

### Statistical analysis

The primary objective was to quantify how model behavior changed as informational content was removed and when counterfactual results created a direct conflict between the provided abstract and the likely “remembered” trial outcome. Performance for the baseline, title-only, and DOI-only conditions was summarized using F1 score (and accuracy) against the original ground-truth label. For the counterfactual condition, the expected label was defined as the inverse of the original ground truth, and performance was evaluated against this counterfactual target. In addition to per-condition performance, we quantified counterfactual sensitivity by calculating the proportion of trials for which the predicted label differed between the baseline and counterfactual inputs (flip rate), and we summarized invalid-format outputs per model and condition. For the calculation of the F1 score, only valid, i.e. correctly formatted predictions were considered. 95% confidence intervals were estimated using normal approximation intervals.

### Ethical consideration

This study used publicly available abstracts from published clinical trials and contained no patient-level information. Therefore, ethics approval was not required.

## Results

Results for all models and input conditions are provided in Table 1. Across conditions, models adhered closely to the required single-token response format. Valid prediction rates ranged from 97.2% to 100.0% depending on model and condition (Figure 1). GPT-5.2 returned 100% valid predictions across all but one condition. Minor format deviations were observed mainly for Gemini (e.g., 97.2% valid in the DOI-only condition and 97.6% in counterfactual settings), while Claude remained near-ceiling (99.2 - 100.0%) across all conditions.

**Table 1.** Performance of GPT-5.2, Gemini 3 Flash, and Claude Opus 4.5 under different conditions. *95% CI = 95% Confidence Interval*

| Condition | Model | Valid predictions [%] | Accuracy (95% CI) | Sensitivity (95% CI) | Specificity (95% CI) | F1 Score (95% CI) |
| --- | --- | --- | --- | --- | --- | --- |
| Baseline | GPT-5.2 | 100.0 | 0.96 (0.94-0.99) | 0.97 (0.94-1.00) | 0.96 (0.92-1.00) | 0.96 (0.94-0.99) |
|  | Gemini 3 Flash | 99.2 | 0.96 (0.94-0.99) | 0.96 (0.93-0.99) | 0.97 (0.94-1.00) | 0.96 (0.94-0.99) |
|  | Claude Opus 4.5 | 100.0 | 0.97 (0.95-0.99) | 0.97 (0.94-1.00) | 0.98 (0.95-1.00) | 0.97 (0.95-0.99) |
| Title only | GPT-5.2 | 100.0 | 0.82 (0.78-0.87) | 0.82 (0.75-0.88) | 0.84 (0.77-0.91) | 0.82 (0.77-0.87) |
|  | Gemini 3 Flash | 99.2 | 0.88 (0.84-0.92) | 0.88 (0.83-0.93) | 0.88 (0.81-0.94) | 0.88 (0.84-0.92) |
|  | Claude Opus 4.5 | 100.0 | 0.79 (0.74-0.84) | 0.79 (0.73-0.86) | 0.78 (0.70-0.86) | 0.78 (0.73-0.83) |
| DOI only | GPT-5.2 | 100.0 | 0.67 (0.61-0.73) | 0.76 (0.69-0.83) | 0.55 (0.45-0.64) | 0.66 (0.60-0.71) |
|  | Gemini 3 Flash | 97.2 | 0.63 (0.57-0.69) | 0.55 (0.47-0.63) | 0.75 (0.67-0.84) | 0.63 (0.57-0.69) |
|  | Claude Opus 4.5 | 99.6 | 0.63 (0.57-0.69) | 0.63 (0.55-0.71) | 0.63 (0.54-0.73) | 0.63 (0.57-0.69) |
| Counterfactual | GPT-5.2 | 99.2 | 0.99 (0.98-1.00) | 0.99 (0.97-1.00) | 0.99 (0.98-1.00) | 0.99 (0.98-1.00) |
|  | Gemini 3 Flash | 97.6 | 0.98 (0.96-1.00) | 0.96 (0.92-1.00) | 0.99 (0.98-1.00) | 0.98 (0.96-1.00) |
|  | Claude Opus 4.5 | 99.2 | 0.96 (0.94-0.99) | 0.92 (0.87-0.97) | 0.99 (0.98-1.00) | 0.96 (0.94-0.99) |
| Counterfactual + DOI | GPT-5.2 | 100.0 | 0.99 (0.97-1.00) | 0.99 (0.97-1.00) | 0.99 (0.97-1.00) | 0.99 (0.97-1.00) |
|  | Gemini 3 Flash | 97.6 | 0.97 (0.94-0.99) | 0.92 (0.87-0.97) | 1.00 (1.00-1.00) | 0.97 (0.94-0.99) |
|  | Claude Opus 4.5 | 99.2 | 0.95 (0.92-0.98) | 0.90 (0.84-0.96) | 0.98 (0.96-1.00) | 0.95 (0.92-0.97) |

**Figure 1.**
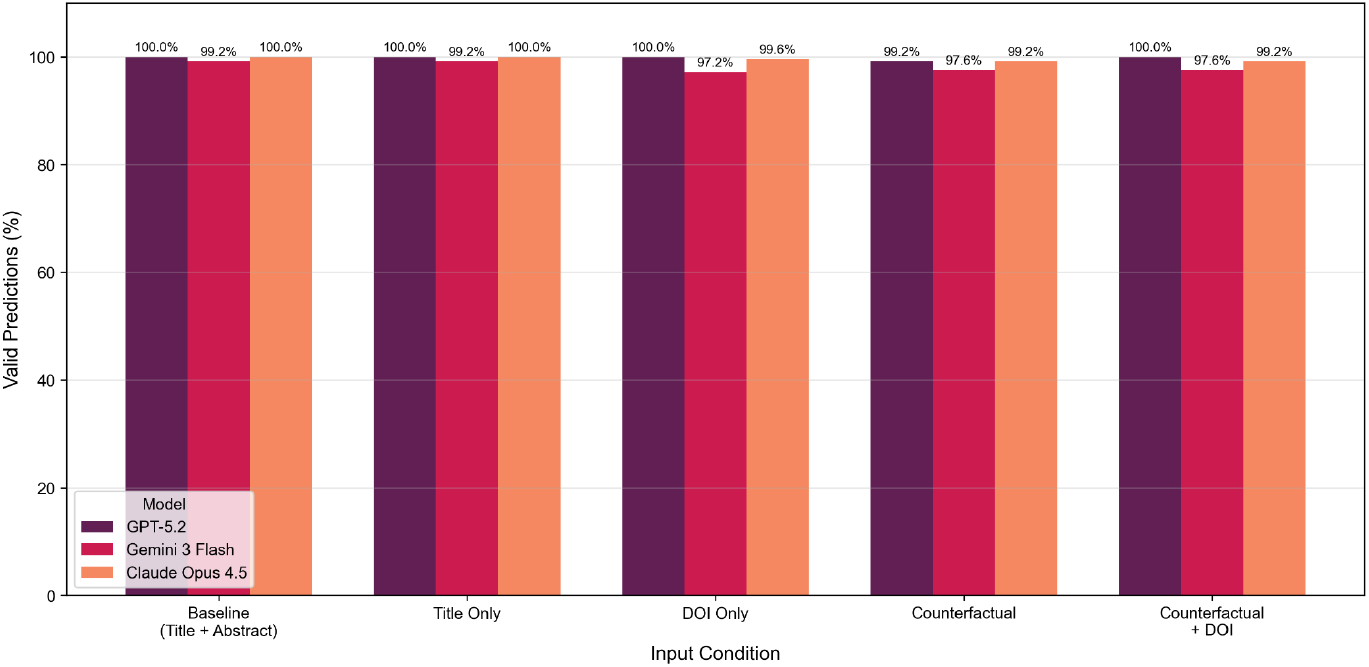
Valid prediction rates of GPT-5.2, Gemini 3 Flash, and Claude Opus 4.5 under different conditions.

The confusion matrices for all models and conditions are provided in Figure 2. When provided with the full title + abstract, all three model families achieved high and tightly clustered classification performance (accuracy: 0.96 - 0.97, F1 Score: 0.96 - 0.97, Table 1). Sensitivity and specificity were similarly high (sensitivity: 0.96 - 0.97, specificity: 0.96 - 0.98), indicating balanced performance for positive and negative trials when the complete abstract text was available.

**Figure 2.**
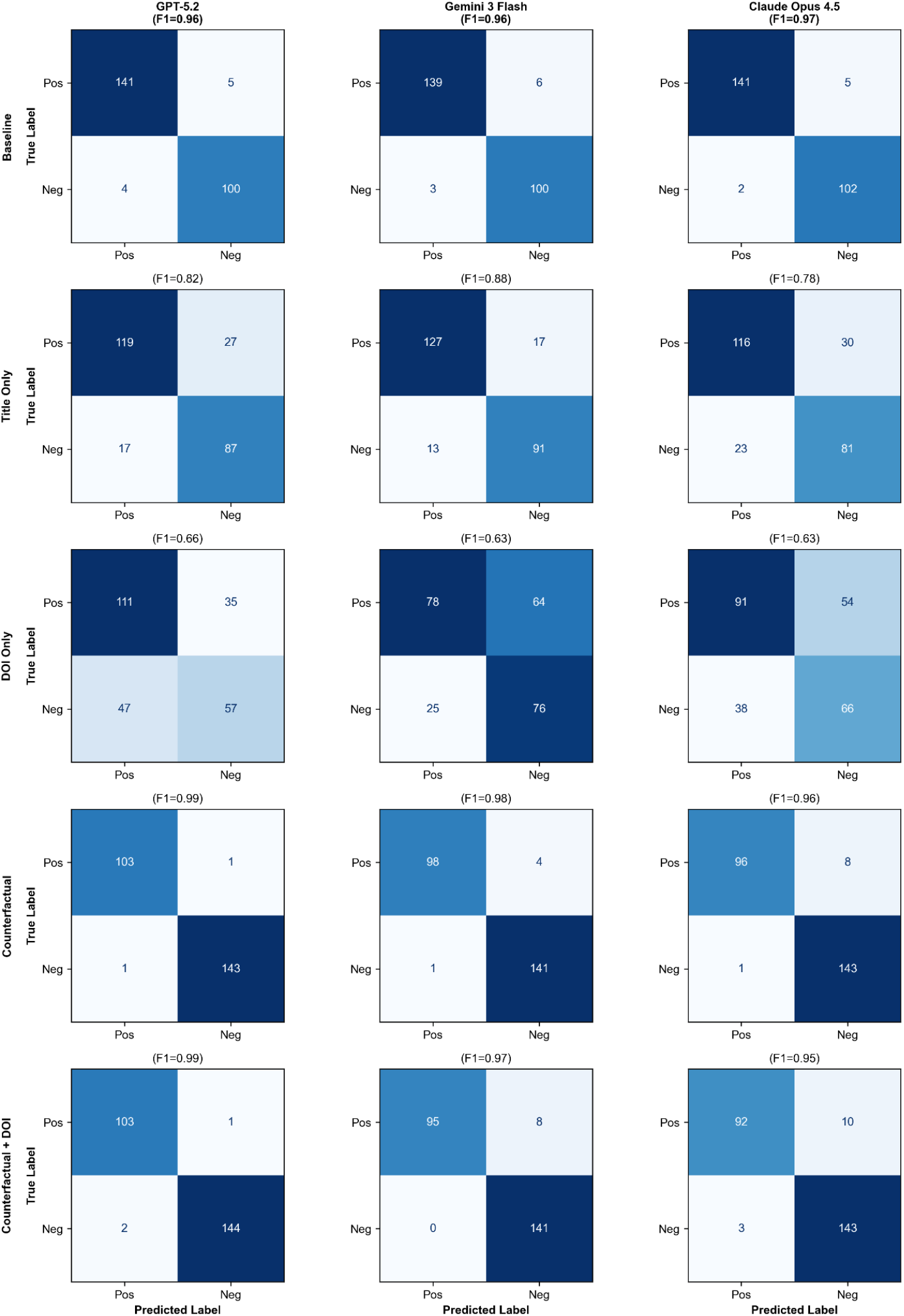
Confusion matrices of GPT-5.2, Gemini 3 Flash, and Claude Opus 4.5 under different conditions.

Removing abstract content reduced performance in a stepwise manner. Under the title-only condition, accuracy fell to 0.79 - 0.88 and the F1 Score to 0.78 - 0.88, with Gemini 3 Flash performing best (accuracy and F1 Score: 0.88) and Claude Opus 4.5 lowest (accuracy: 0.79, F1 Score: 0.78). Sensitivity and specificity remained broadly comparable within each model (GPT: 0.82 vs 0.84, Gemini: 0.88 vs 0.88, Claude: 0.79 vs 0.78), suggesting that performance degradation with titles alone was not driven solely by one-sided misclassification. With DOI-only inputs, performance decreased further for all models (accuracy: 0.63 - 0.67, F1 Score: 0.63 - 0.66,). Error profiles diverged by model: GPT-5.2 showed higher sensitivity than specificity (0.76 vs 0.55), whereas Gemini showed the reverse pattern (0.55 vs 0.75), and Claude remained relatively symmetric (0.63 vs 0.63). The F1 Scores for all models and conditions are visualized in Figure 3.

**Figure 3.**
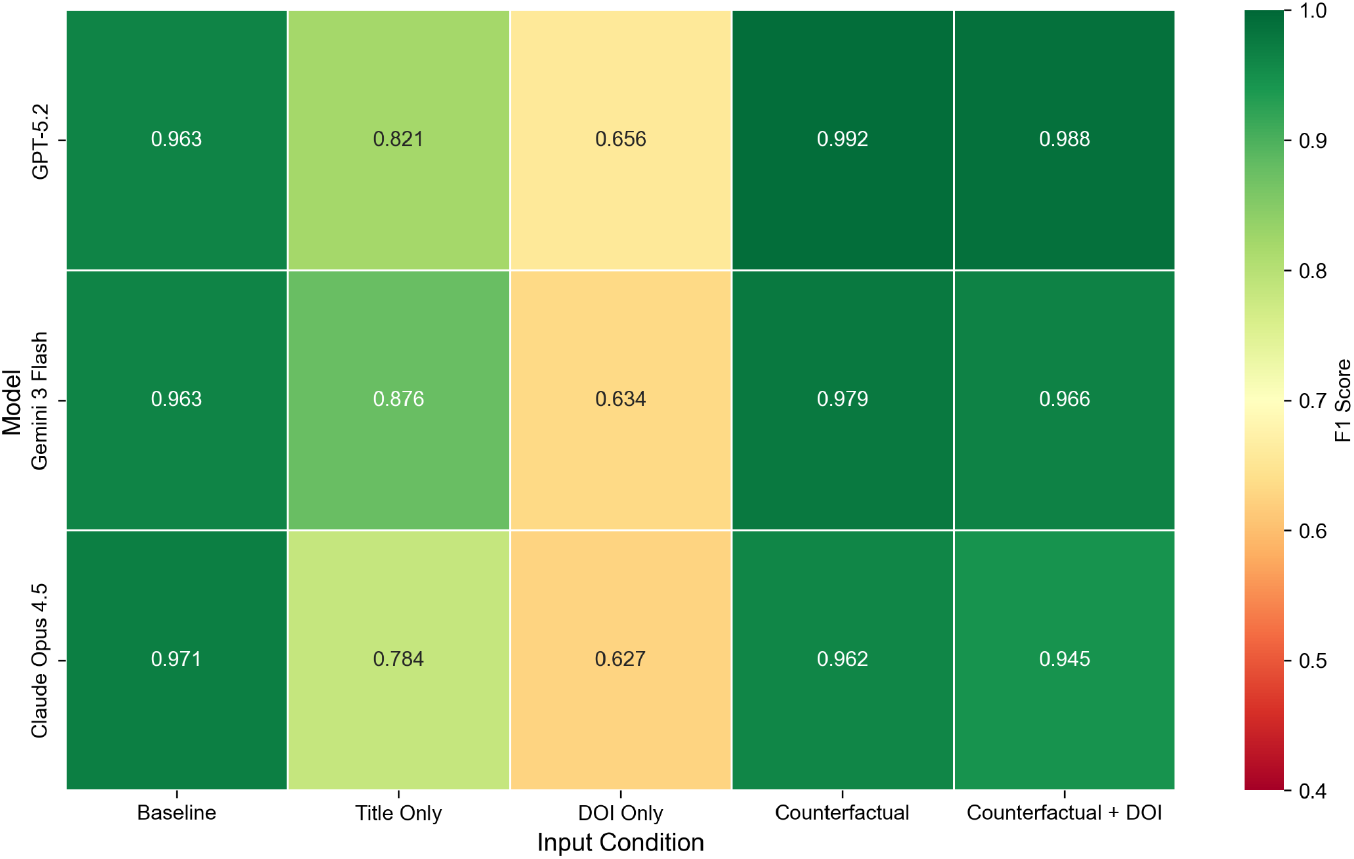
Heatmap of F1 Scores of GPT-5.2, Gemini 3 Flash, and Claude Opus 4.5 under different conditions.

To generate counterfactual abstracts, edits were concentrated in outcome-bearing portions of the text. Across 250/250 trials, the results and conclusion sections required modification (100.0% each). In contrast, edits were rarely required in the Title (13/250, 5.2%) or Methods (4/250, 1.6%), and the Introduction did not require modification (0/250, 0.0%).

Against the inverted ground-truth labels, models followed the counterfactual evidence with near-ceiling performance. In the counterfactual condition, accuracy and F1 Score ranged from 0.96 - 0.99 (GPT: 0.99, Gemini: 0.98, Claude: 0.96). Specificity remained high (0.99 for all three models), while Claude’s reduction relative to GPT/Gemini was driven mainly by lower sensitivity (0.92, 95% CI 0.87 - 0.97).

When the original identifier was reintroduced (counterfactual + DOI), performance changed minimally for GPT-5.2 (accuracy and F1 Score: 0.99) but decreased modestly for Gemini (accuracy and F1 Score: 0.97) and Claude (accuracy and F1 Score: 0.95). Notably, Gemini maintained perfect specificity (1.00) in this condition (95% CI 1.00 - 1.00) alongside reduced sensitivity (0.92), whereas Claude showed a larger sensitivity drop (0.90) with specificity still high (0.98).

## Discussion

All three commercial model families achieved high performance when given full title + abstract information (accuracy and F1 Score clustered around 0.96 - 0.97), with only minor rates of invalid-format outputs. Performance degraded stepwise as informational content was removed: With title-only inputs, accuracy dropped to ∼0.79 - 0.88, and with DOI-only inputs it fell further to ∼0.63 - 0.67. In contrast, when the abstract’s primary-outcome statement was counterfactually flipped, models followed the altered evidence and achieved near-ceiling performance against the inverted labels (accuracy and F1 Score ∼0.96 - 0.99). Reintroducing the real DOI alongside the counterfactual abstract minimally affected GPT-5.2 but led to modest performance reductions for Gemini and Claude, primarily via reduced sensitivity, suggesting occasional identifier-driven interference when DOI and results text conflicted.

Interpreting these results in the context of prior work, the strong baseline performance is consistent with broader evidence that LLMs can perform competitively on biomedical language tasks.^1,3^ However, high task performance alone does not establish that a model is “reading” the provided document rather than drawing on parametric knowledge, an issue long discussed in the framing of language models as implicit knowledge bases.^4^ The stepwise ablation results speak directly to that distinction. Title-only performance well above chance can plausibly arise from a mixture of (i) genuine, weakly informative cues in titles (e.g., disease setting, intervention class, endpoint hints), and (ii) recognition of well-known trials from pretraining exposure. DOI-only inputs are especially diagnostic because the DOI string contains minimal semantic content for endpoint attainment. Above-chance performance under DOI-only conditions is therefore most consistent with identifier-triggered recall/recognition or correlated metadata effects (e.g., publisher/journal patterns).

However, the counterfactual results indicate that the models don’t allow their reasoning to be overridden by their internal memory: When the outcome-bearing sentences in the abstract were minimally edited to flip the primary endpoint conclusion, models overwhelmingly produced the inverted label, indicating sensitivity to the supplied text even when that text contradicted what a model might have “seen before.” At the same time, the small but consistent decrement for some models when the real DOI was appended to the counterfactual abstract suggests that identifiers can still exert a measurable pull on the model’s decision, echoing findings that models do not integrate context and prior knowledge uniformly and can privilege priors when they are “familiar” with an entity.^9^ It is important to emphasize that our counterfactual flips created direct, high-salience evidence conflicts. In more realistic settings, users may supply incomplete or subtly contradictory evidence, where LLMs can struggle with conflict resolution in biomedical contexts.^12^ Taken together, the findings support a nuanced conclusion: The models can robustly follow explicit outcome statements in abstracts (strong “reading” behavior under clear evidence), yet still display some degree of susceptibility to identifier-driven priors when content is sparse or when identifiers are introduced.

Several strengths support the interpretability and practical relevance of these findings. First, the design operationalizes “read versus remember” using deterministic, logged perturbations (progressive content removal plus counterfactual outcome edits), which makes the diagnostic logic transparent and reproducible. Second, the evaluation uses a corpus of real oncology RCT abstracts spanning multiple high-impact journals and many publication years, aligning the test distribution with how LLMs are used in evidence synthesis and clinical-research workflows rather than relying solely on synthetic benchmarks. Third, the single-token output constraint (POSITIVE/NEGATIVE) reduces ambiguity in scoring and minimizes the confounding role of verbose explanations, while the high valid-output rates indicate that the API-based setup was stable across conditions. Fourth, comparing multiple commercial model families under default settings improves external validity for typical end-user deployment, where practitioners rarely tune decoding parameters or implement specialized grounding interventions.

Several limitations should temper overgeneralization. First, the study focuses on a single binary task (primary endpoint met vs not met) within oncology RCTs. Reading–remembering dynamics may differ for tasks requiring finer-grained extraction, multi-label judgments, or synthesis across multiple documents. Second, DOI-only above-chance performance could partly reflect indirect correlations encoded in DOI structure (publisher prefix, journal family, year) rather than true memorized recall of the trial outcome. Additional controls (e.g., DOI-shuffling across papers, synthetic DOIs matched on prefix/year) would help isolate the mechanism. Third, counterfactual editing was designed to be minimal, but any manual or rule-based editing procedure risks introducing artifacts (lexical cues, unnatural phrasing, or consistency breaks) that models could exploit. Even though edits were concentrated where outcomes are stated, future work could quantify detectability of edits or use blinded human review to ensure counterfactual naturalness. Fourth, models were queried via vendor APIs with default settings and without a fixed seed. While this reflects realistic usage, it limits analysis of variance due to decoding stochasticity and complicates strict reproducibility across future model snapshots. Finally, we did not evaluate model calibration, abstention behavior, or uncertainty reporting - properties that may be crucial when deploying such systems in safety-critical evidence workflows.

Future research could try to strengthen causal attribution of identifier effects by adding negative controls: Swapping DOIs between trials, adding semantically irrelevant identifiers, or using “matched” synthetic DOIs, which would help separate true memorized mapping from metadata correlations. A second direction is to broaden task coverage: Applying the same progressive-ablation and counterfactual-conflict framework to other biomedical NLP tasks (PICO extraction, effect direction and magnitude, toxicity endpoints, comparative effectiveness statements) would test whether strong counterfactual sensitivity generalizes beyond this relatively explicit classification problem. A third direction is temporal and contamination-robust evaluation: Constructing time-split test sets consisting of trials published after known model training cutoffs (or using newly published/embargoed material where feasible) would reduce the plausibility of parametric recall and better isolate reading-based competence

In conclusion, this study suggests that contemporary commercial LLMs can faithfully use the information in oncology RCT abstracts when outcome statements are explicitly present, as shown by near-ceiling performance on counterfactual abstracts that directly contradict the original results. At the same time, above-chance performance with titles - and especially with DOI-only inputs – indicates that identifiers can carry predictive signal for these models, consistent with some degree of recognition or learned association that is independent of the provided text. The modest degradation observed when reintroducing real DOIs into counterfactual abstracts further implies that identifier-triggered priors can occasionally compete with textual evidence.

## Data Availability

All data and code used to obtain this study's results have been uploaded to https://github.com/windisch-paul/llm_memory.

https://github.com/windisch-paul/llm_memory

